## Supplement 2: STROBE Checklist for "Community Learning Ledgers for Cancer Navigation in Small Island Developing States"

### eChecklist. STROBE reporting map

**Community Learning Ledgers for Cancer Navigation in Small Island Developing States** — reporting checklist for observational studies (STROBE, cohort/cross-sectional). Each item maps to the manuscript section that satisfies it; items not applicable to a retrospective ledger study state why.

| # | STROBE item | Where addressed |
|---|---|---|
| 1 | Title and abstract: design named; balanced summary | Title page; structured Abstract (design named in Design, Setting, and Participants) |
| 2 | Background/rationale | Introduction |
| 3 | Objectives | Abstract (Objective); Introduction, closing paragraph |
| 4 | Study design | Methods — Study Design and Oversight |
| 5 | Setting: locations, dates, periods | Methods — Study Design; Results (March 2 – June 1, 2026; ten Caribbean jurisdictions) |
| 6 | Participants: eligibility, sources, selection | Methods — Community Learning Ledger (all sessions; no registration; anonymized at capture) |
| 7 | Variables: outcomes, exposures, definitions | Methods — Thematic Classification (codebook, eTable 2); Multi-Arm Analysis (three binary outcomes) |
| 8 | Data sources/measurement | Methods — Community Learning Ledger (three-field schema); Multi-Arm Analysis (verbatim submission, deterministic scoring) |
| 9 | Bias | Limitations (ecological benchmark; retrospective design; single-region deployment) |
| 10 | Study size | Results (N=207 sessions; 28-query Cohort A; 14-query Cohort B) |
| 11 | Quantitative variables | Methods — Statistical Analysis (binary outcomes; exact tests) |
| 12 | Statistical methods | Methods — Statistical Analysis (McNemar exact test, Holm correction, exact binomial CIs) |
| 13 | Participants: numbers at each stage | Results (207 sessions; 179 substantive; 21 non-substantive; 7 adversarial) |
| 14 | Descriptive data | Results — Community Learning Accumulation; eTable 9 |
| 15 | Outcome data | Results — Multi-Arm Comparative Navigation Analysis; Tables 1–2; eTable 4 |
| 16 | Main results: estimates, precision | Results (percentages with 95% CIs; paired comparisons with P values) |
| 17 | Other analyses | Results — Secondary Outcomes; Emergent Safety Finding; CARIVIVA corroboration |
| 18 | Key results summary | Discussion, opening; Conclusions |
| 19 | Limitations | Addressed in the dedicated Limitations section |
| 20 | Interpretation | Discussion |

| # | STROBE item | Where addressed |
|---|---|---|
| 21 | Generalisability | Discussion (SIDS generalization; Malta comparison, eAppendix B) |
| 22 | Funding | Stated in its own section: no external grants; CANONIC Foundation infrastructure support; funder role declared |

The study is a retrospective observational analysis of an anonymized community ledger with a prospective comparative arm; consent items and follow-up items specific to interventional cohorts do not apply — the platform collects no identifiers and the study protocol is filed for exempt determination under 45 CFR 46.104(d)(4)(ii).
